# In-Hospital Survival of Adults with HIV-Associated Cryptococcal Meningitis in Tanzania: A Retrospective Comparison of Amphotericin B-based Regimen and Fluconazole Monotherapy

**DOI:** 10.1101/2025.09.07.25335283

**Authors:** Mlela Msongela, George Musiba, Juma A. Mohamed, Alphonce I. Marealle, Manase Kilonzi, Ritah F. Mutagonda

## Abstract

**Objective:** This study aimed to examine the prevalence, treatment practices, outcomes, and factors associated with in-hospital survival (discharged alive) among cryptococcal meningitis (CM) patients living with HIV in Tanzania.

**Methods:** This hospital-based cross-sectional study retrospectively reviewed records of people living with HIV (PLHIV) admitted to medical wards at Dodoma and Singida Regional Referral Hospitals in Tanzania from July 2019 to June 2024. Data on socio-demographics, antiretroviral therapy (ART) status, CD4 count, CM status, treatment, and outcomes were extracted using a standardised data collection tool. The primary outcome was in-hospital survival (discharged alive vs died). Descriptive statistics summarised patient characteristics, and modified Poisson regression with robust variance estimated adjusted risk ratios (aRR) for factors associated with being discharged alive.

**Results:** A total of 561 PLHIV records were reviewed. Of these, 288 (51.5%) were aged 36–55 years, 309 (55.1%) were female, and 435 (77.5%) were in WHO clinical stage IV. Overall, 159 (28.3%) patients had CM, of whom 88 (55.3%) received fluconazole monotherapy. In-hospital mortality among CM patients was 65 (46.5%). Discharge alive occurred in 61/73 (83.6%) of those on amphotericin B–based regimens versus 13/66 (19.7%) on fluconazole monotherapy. Patients treated with amphotericin B–based regimens were four times more likely to be discharged alive compared to those on fluconazole monotherapy (aPR = 4.19, 95% CI: 2.46–7.16, p < 0.001).

**Conclusion:** CM remains a major opportunistic infection among PLHIV, with most patients managed using fluconazole monotherapy. In-hospital survival was significantly higher with amphotericin B–based regimens, highlighting the need to align practice with guideline recommendations. Further qualitative research is warranted to explore barriers to implementing recommended CM treatment.

## I. Introduction

Cryptococcus meningitis (CM) is the most common opportunistic infection and remains the leading cause of mortality in people living with HIV (PLHIV) (1). Globally, CM is estimated to cause 350,000 to 1.5 million new cases annually, causing a mortality of approximately 181,000, with around 75% of deaths being reported in sub-Saharan Africa. In Africa prevalence of CM in PLHIV ranges from 5.1% to 33%, with the majority of these cases and mortality occurring in SSA. For instance, studies conducted in South Africa, Ethiopia, Uganda, and Kenya reported a prevalence of 7%,7% 11%, and 33%Respectively (2,3). Also, a study conducted in Dar es Salaam, Tanzania, reported a prevalence of 11.5% and an in-hospital mortality rate range of 50% to 70% (2,4–7).

Cryptococcal meningitis (CM) can be treated with various antifungal agents, including fluconazole, voriconazole, amphotericin B, flucytosine, and Posaconazole. However, susceptibility to these agents varies between *Cryptococcus gattii* and *Cryptococcus neoformans*, as well as across different global regions. Historically, CM treatment relied on fluconazole monotherapy at 800 to 1,200 mg daily doses. Over time, this approach has been associated with the emergence of antifungal resistance, limited fungicidal activity, and high mortality rates (8–11). In response, the World Health Organisation (WHO) updated guidance to recommend amphotericin B–based induction, specifically single-dose liposomal amphotericin B (10 mg/kg) plus 14 days of flucytosine and fluconazole, which has demonstrated non- inferior survival, improved renal safety, and acceptable tolerability when appropriately monitored (12–14).

Amphotericin B-based regimen is reported to have more rapid clearance of *Cryptococcus* from both serum and cerebrospinal fluid (CSF) cultures. This regimen is highly fungicidal and significantly reduces in-hospital mortality compared to fluconazole monotherapy (10,11,14–16). Guidelines recommend a single high dose (10 mg/kg) of liposomal amphotericin B, administered in combination with 14 days of flucytosine and fluconazole. This approach has demonstrated non-inferior survival outcomes, improved renal safety, and acceptable tolerability when patients are closely monitored (13).

Tanzania, as a member state of the WHO, updated its national HIV treatment guidelines to align with the recommendations, which include the use of amphotericin B-based regimen for the treatment of CM in PLHIV. Despite this recommendation, clinical information on the implementation and outcomes of this triple therapy remains limited. A previous descriptive study conducted in Dar es Salaam reported that only 22.8% of 45 patients with CM received triple therapy, and favourable outcomes were primarily observed among those treated with this regimen (4). However, this study was limited by its small sample size and did not evaluate the comparative likelihood of survival (i.e., being discharged alive) between patients treated with amphotericin B-based regimen versus those who received fluconazole monotherapy. To address this gap, we assessed, among PLHIV admitted to two Tanzanian regional referral hospitals, the prevalence of CM, treatment modalities used, in-hospital outcomes, and factors associated with being discharged alive, with a primary comparison between amphotericin B–based regimens and fluconazole monotherapy.

## II. METHODS

### A. Study Design and Setting

This was a hospital-based, cross-sectional study involving retrospective data abstraction. Data were collected from patient records spanning July 2019 to June 2024. The abstraction process was conducted between March and April 2025 at Dodoma Regional Referral Hospital (DRRH) and Singida Regional Referral Hospital (SRRH) in Tanzania. Both facilities are secondary-level referral centres for the central zone, each hosting a Care and Treatment Centre (CTC) for PLHIV and internal medicine wards providing inpatient care (17).

### B. Study population

The source population comprised all PLHIV admitted to the medical wards of DRRH and SRRH within the study period. From this frame, we identified the CM cohort as those with a clinician diagnosis of cryptococcal meningitis documented in the chart and/or supporting laboratory evidence (e.g., positive CSF or serum cryptococcal antigen, India ink, or culture), recorded during the index admission.

### C. Sample size and calculation

All eligible records within the study window were included. For planning the precision of descriptive estimates (CM prevalence among PLHIV admissions), we calculated a minimum sample size using the Leslie Kish formula for cross-sectional studies, N=Z2×P(1−P)/d2N = Z^2 \times P(1-P) / d^2N=Z2×P(1−P)/d2 (18), with Z=1.96Z = 1.96Z=1.96 (95% confidence), P=0.12P = 0.12P=0.12 based on a prior Dar es Salaam study (4), and d=0.05d = 0.05d=0.05. This yielded N=163N = 163N=163; allowing 20% for incomplete/missing records, increased the target to 204. The final cohort exceeded this minimum.

### D. Data collection procedure

Data were collected from patient files and electronic hospital management systems using a structured abstraction tool adapted from a previous study conducted in Dar es Salaam, Tanzania (4). The tool was used to extract relevant information from patient medical records, including age, sex, marital status, health insurance status, occupation, antiretroviral therapy (ART) status, CD4 count, CM status, treatment modality, and treatment outcomes.

The data abstraction process was led by the principal investigator, a second-year Master’s student in the Hospital and Clinical Pharmacy program, in collaboration with two trained research assistants. Each assistant held a Bachelor of Pharmacy degree and was stationed at one of the selected RRHs. Before data collection, the research assistants were trained on the study objectives and received a comprehensive orientation on the data abstraction tool.

To ensure consistency and data quality, the principal investigator supervised the initial days of abstraction at each site and conducted ongoing consistency checks and logic/range checks to resolve discrepancies.

### E. Data Analysis

Data was initially entered into Microsoft Excel and subsequently imported into IBM SPSS Statistics version 27 for cleaning and analysis. Categorical variables were summarised using frequencies and percentages. Bivariate associations with the primary outcome were screened using Pearson’s chi-square; variables with p≤0.20 entered multivariable models alongside a priori confounders (site and calendar period). For the primary effect estimate, we fitted modified Poisson regression with robust variance to obtain adjusted risk ratios (aRRs) and 95% confidence intervals for being discharged alive comparing amphotericin B–based therapy versus fluconazole monotherapy. We assessed multicollinearity and retained variables with clinical relevance and/or statistical support. Mediator candidates (e.g., completion of induction) were not included in the primary model.

Twenty (20) CM patients who received treatment were excluded from this analysis, as their treatment outcomes could not be determined due to missing or incomplete records (i.e., loss to follow-up). Therefore, the regression analyses were conducted on the remaining patients with clearly documented outcomes, ensuring the validity of the associations identified.

### F. Ethical considerations

Ethical approval was obtained from the MUHAS Research and Ethics Committee (Ref. DA.282/298/01.C/2549). Hospital administrations at DRRH and SRRH granted permission for data access. No personal identifiers (e.g., names, hospital numbers) were abstracted. All procedures complied with institutional policies and the Declaration of Helsinki.

## III. Results

### A. Socio-demographic and Clinical Characteristics

A total of 561 patient medical records were reviewed. The majority of patients, 289 (51.5%), were aged between 36 and 55 years, and females accounted for 309 (55.1%). Most patients, 112 (20.0%), had active health insurance, and 409 (72.9%) were known to be HIV-positive. Additionally, 435 (77.5%) were classified as WHO clinical stage IV, while 361 (64.3%) had a history of antiretroviral therapy (ART) defaulter. A history of ART default was recorded in 361 (64.3%). CD4 <200 cells/mm³ was documented in 505 (90.0%). Approximately half had at least one comorbidity (274; 48.8%), *Table 1*.

**Table 1:** Characteristics of Participants in the Study (N=561)

| Variable | Frequenc<br>y | Percentag<br>e |
| --- | --- | --- |
| **Year of Treatment** |  |  |
| 2019-2020 | 82 | 14.6% |
| 2021-2022 | 127 | 22.6% |
| 2023-2024 | 352 | 62.8% |
| **Sex** |  |  |
| Female | 309 | 55.1% |
| Male | 252 | 44.9% |
| **Age in years (Mean: 44.86, Std: 13.68)** |  |  |
| below 35 | 150 | 26.8% |
| 36 to 55 | 289 | 51.5% |
| Above 55 | 122 | 21.7% |
| **Insurance Status** |  |  |
| Insured | 112 | 20.0% |
| Not Ensured | 449 | 80.0% |
| **Marital Status** |  |  |
| Divorced | 48 | 8.5% |
| Married | 308 | 54.9% |
| Separated | 38 | 6.8% |
| Single | 114 | 20.3% |
| Widowed | 53 | 9.4% |
| **Occupation** |  |  |
| Employed | 117 | 20.9% |
| Self Employed | 283 | 50.4% |
| Student | 24 | 4.3% |
| Unemployed | 137 | 24.4% |
| **CD4 Count [Median: 182.81 (59.75 – 742.00)]** |  |  |
| $\geq 200$ cells/mm<sup>3</sup> | 56 | 10.0% |
| < 200 cells/mm<sup>3</sup> | 505 | 90.0% |
| **HIV Diagnosis** |  |  |
| Known HIV Patient at CM treatment | 409 | 72.9% |
| Newly Diagnosed at CM treatment | 152 | 27.1% |
| **WHO Clinical Stage** |  |  |
| Stage I | 7 | 1.2% |
| Stage II | 27 | 4.8% |
| Stage III | 92 | 16.4% |
| Stage IV | 435 | 77.5% |
| **ART Defaulter** |  |  |
| Yes | 361 | 64.3% |
| No | 200 | 35.7% |
| **Comorbidity** |  |  |
| Yes | 274 | 48.8% |
| No | 287 | 51.2% |
| **Comorbidities (N = 274)** |  |  |
| Anaemia | 8 | 1.4% |
| Chronic Kidney Disease | 6 | 1.1% |
| Central Nervous System disorders | 14 | 2.5% |
| Cardiovascular System disorder (Hypertension, Heart failure) | 47 | 8.4% |
| Diabetes Mellitus Type 2 | 10 | 1.8% |
| Liver Disease | 8 | 1.4% |
| Malignancy | 9 | 1.6% |
| Opportunistic Infections (Toxoplasmosis, Pneumocystis Jirovecii pneumonia, pulmonary tuberculosis, Herpes) | 144 | 25.7% |
| Others | 28 | 5.0% |

### B. Cryptococcal Meningitis Prevalence and Treatment Modalities

Among 561 patients reviewed, 159 (28.3%) were confirmed to have CM. Of these, 71 (44.7%) received an amphotericin B–based regimen, whereby baseline renal function testing was documented for 52 (73.2%), and completion of the induction phase was reported for 48 (67.6%), *Table 2*.

**Table 2:** Cryptococcal Meningitis Prevalence, Treatment Modalities, and Treatment Outcome.

| **Prevalence of Cryptococcal meningitis (N=561)** | **Frequency** | **Percentage** |
| --- | --- | --- |
| Positive | 159 | 28.30% |
| Negative | 402 | 71.70% |
| **Treatment Modality (N=159)** |  |  |
| Fluconazole Monotherapy | 88 | 55.30% |
| Amphotericin B- based Triple Therapy | 71 | 44.70% |
| **Completion of Induction Phase (N=71)** |  |  |
| Yes | 48 | 67.60% |
| No | 23 | 32.40% |
| **Renal Function Tests Done (N=71)** |  |  |
| Yes | 52 | 73.20% |
| No | 19 | 26.80% |
| **Treatment Outcomes (N=139)** |  |  |
| Discharged alive | 65 | 40.9% |
| In-Hospital Mortality | 74 | 46.5% |
| Lost to follow-up | 19 | 11.9% |
| Referred | 1 | 0.7% |

For all CM cases (n=159), 74 (46.5%) were discharged alive and 65 (40.9%) had in-hospital death. The primary analytic set for comparative analyses excluding LTFU and referrals, yielded N=139 CM patients with known outcomes. In this analytic set, discharged alive was 74/139 (53.2%), and in-hospital death was 65/139 (46.8%).

### C. Factors associated with being discharged alive among study participants

In bivariable analyses, female sex and married/separated status were associated with higher crude likelihood of discharge alive; however, these associations were not statistically significant after adjustment. In the multivariable modified Poisson model, receipt of an amphotericin B–based regimen remained strongly associated with discharge alive compared with fluconazole monotherapy (aRR 4.19, 95% CI 2.46–7.16; p<0.001)., *table 3*

**Table 3:**
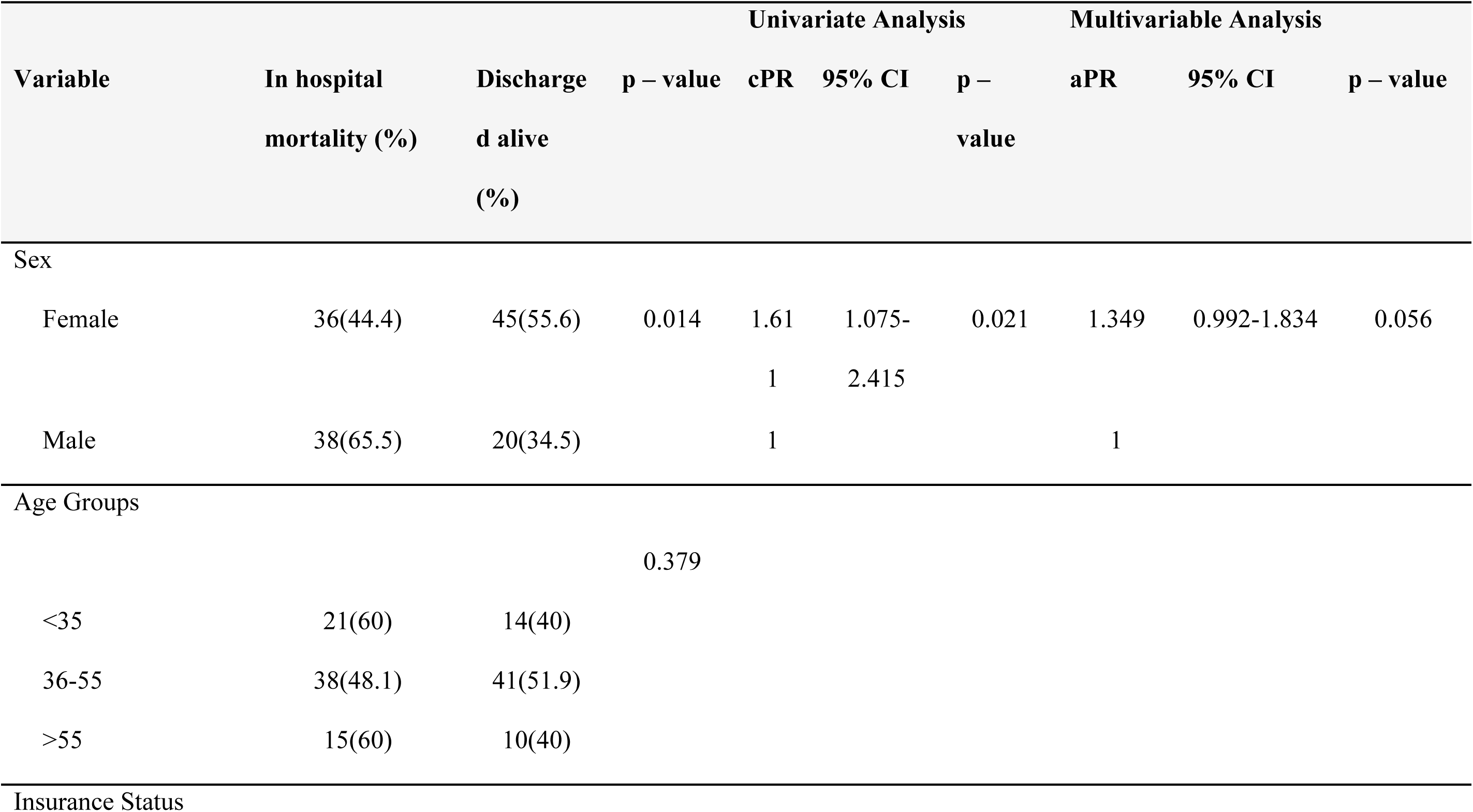
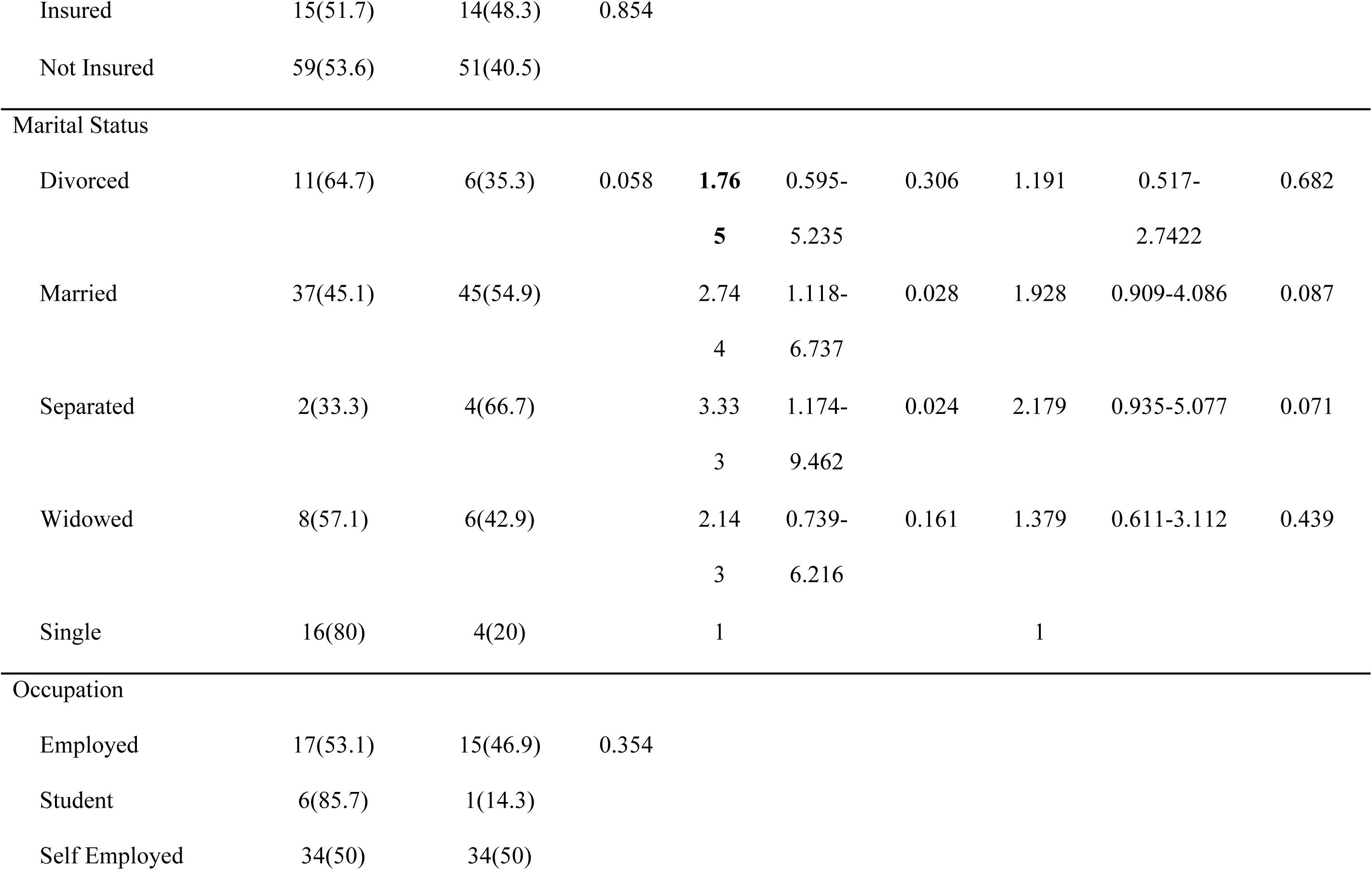
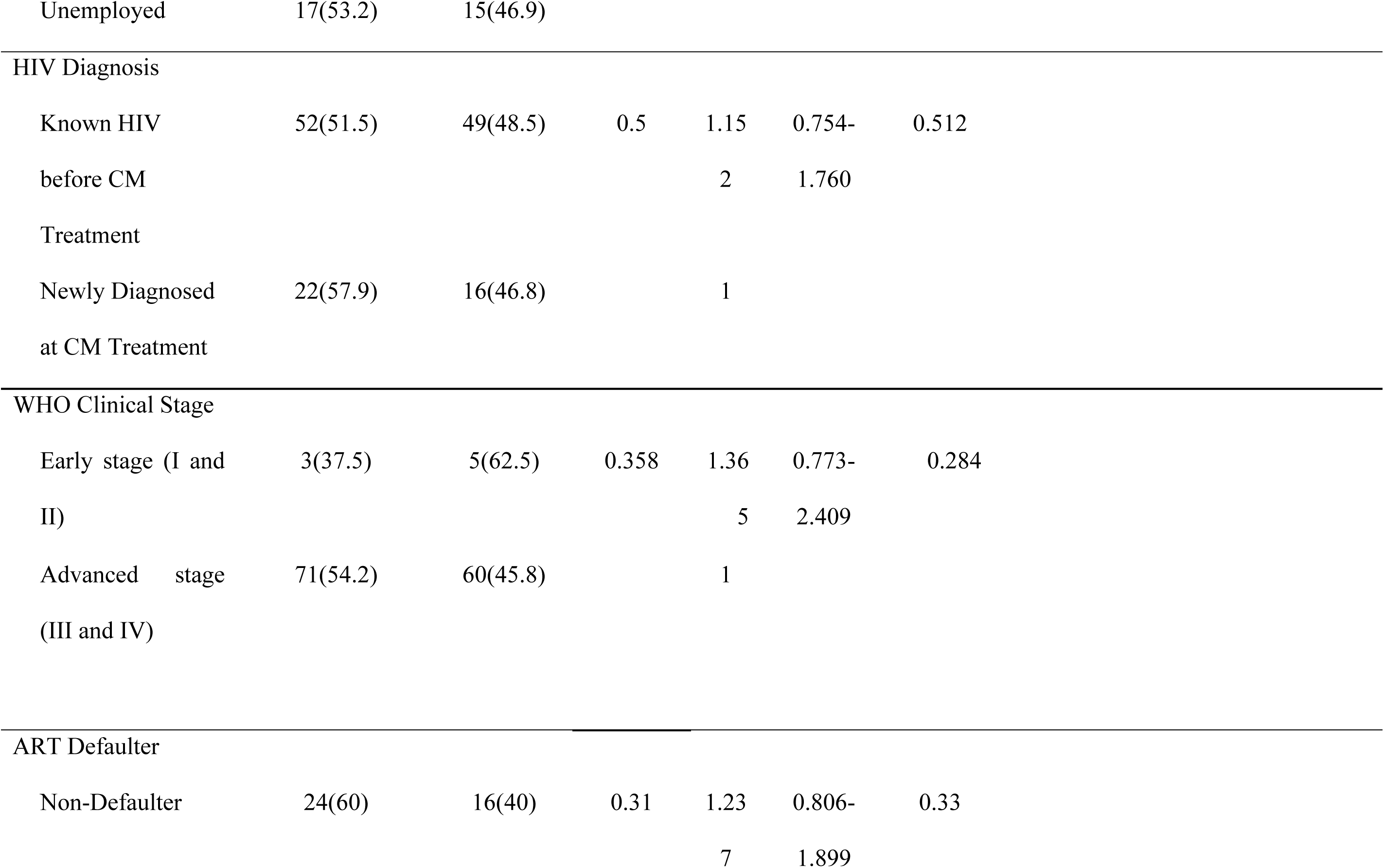
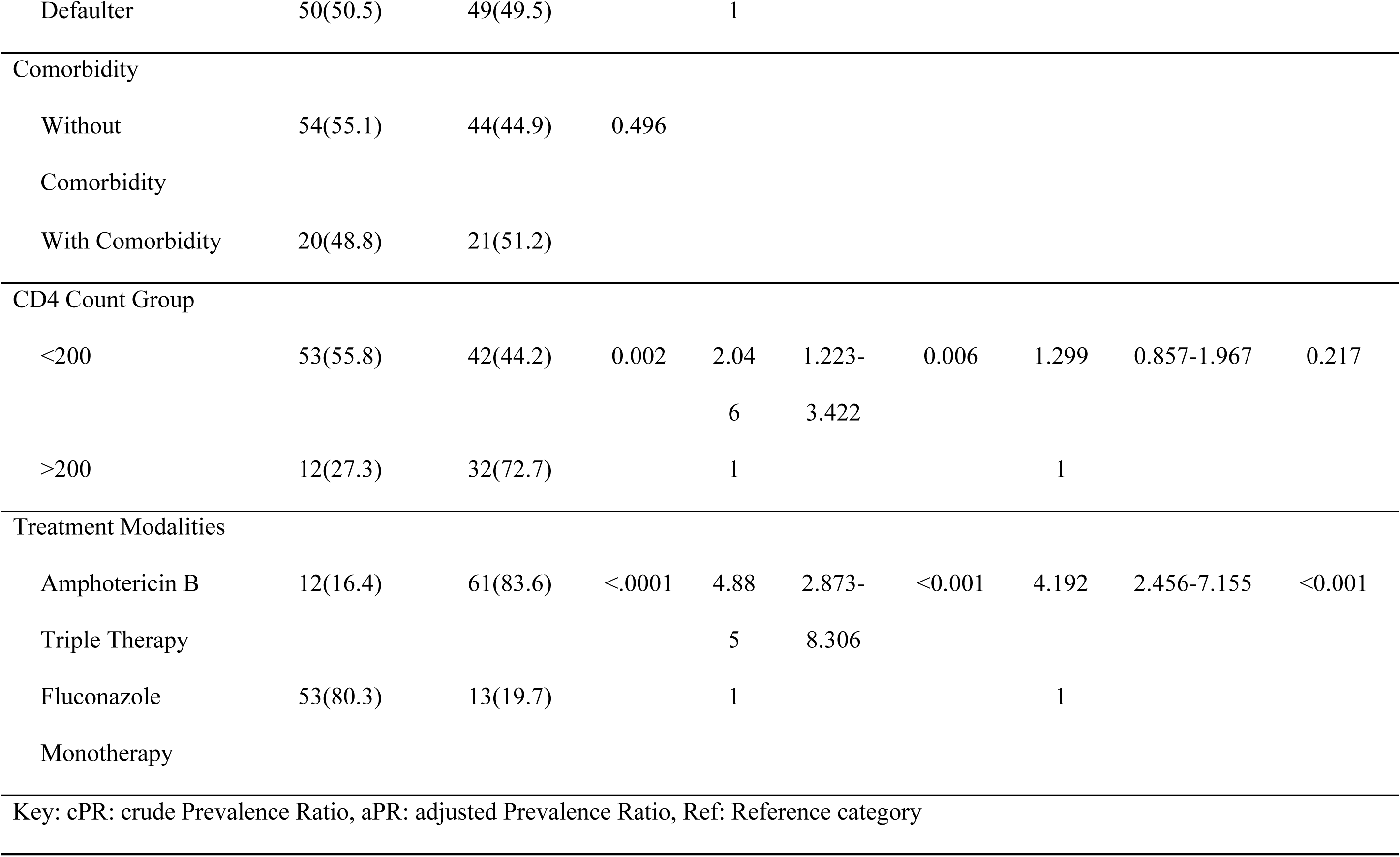
Robust Poisson regression for factors associated with being discharged alive among participants (N = 139)

### IV. Discussion

CM remains a major opportunistic infection among PLHIV, with a prevalence of 28.3% among admitted patients over five years. This study examined the prevalence, treatment practices, outcomes, and determinants of being discharged alive among CM patients living with HIV in Tanzania. In the analytic set, discharge alive occurred in 83.6% of patients receiving amphotericin B–based therapy versus 19.7% on fluconazole monotherapy. Across the full CM cohort (N=159), in-hospital mortality was 46.5% and fluconazole monotherapy remained common (88/159; 55.3%) despite contemporary guidance.

This study demonstrates that CM continues to affect a considerable proportion of PLHIV in regions where the disease is endemic. The prevalence observed in our cohort is comparable to that reported in Kenya (33%), but markedly higher than estimates from previous studies conducted in Dar es Salaam, Tanzania (11.9%), Ethiopia (11.4% and 7.7%), Cameroon (11.2%), and Sierra Leone (8%) (2,4,18–21). These variations are likely attributable to differences in study design, population characteristics, and data sources; some were cross-sectional, prospective, or retrospective, while others focused on patients attending HIV clinics, hospital admissions, or data retrieved from medical records over time (2).

Although the scale-up of effective ART and the adoption of “test-and-treat” strategies have substantially reduced HIV-related morbidity, mortality, and transmission, CM remains a significant and life-threatening opportunistic infection. Our findings underscore the urgent need to strengthen early HIV diagnosis, timely ART initiation, and sustained adherence support, as well as to monitor emerging HIV-1 resistance patterns that may influence opportunistic infection risk. This is consistent with a multicentre study estimating an annual CM burden of approximately 162,500 cases in Africa, largely attributed to delayed ART initiation (6). Supporting this, studies from Rwanda and South Africa reported a lower prevalence of CM, which was strongly associated with earlier ART initiation (22–24).

Our study revealed notable variability in CM treatment, with most patients receiving fluconazole monotherapy. This aligns with findings from a previous study in Dar es Salaam, Tanzania, where more than 80% of CM patients were treated with fluconazole monotherapy (4). Such practice, however, contrasts with both national and WHO guidelines, which recommend amphotericin B–based regimens, preferably a combination of amphotericin B, flucytosine, and fluconazole, or amphotericin B with fluconazole when flucytosine is unavailable (14,25). Process indicators revealed additional implementation gaps whereby baseline renal function testing was documented in 73.2% of amphotericin recipients, and only 67.6% completed the induction phase. Guideline recommendations emphasize renal monitoring due to the drug’s nephrotoxicity, and incomplete induction therapy has been strongly linked to poor outcomes (24). Importantly, our study demonstrated that patients treated with amphotericin B-based regimens were four times more likely to be discharged alive compared to those receiving fluconazole monotherapy. These findings are consistent with evidence from multiple clinical trials showing superior effectiveness of amphotericin B-based therapy, reflected in shorter hospital stays and lower in-hospital mortality (**7,12,26–28**).

The continued reliance on fluconazole monotherapy and suboptimal completion of amphotericin B therapy likely stems from several factors, including limited awareness of treatment guidelines, inadequate provider training in amphotericin B preparation and administration, irregular drug supply, and the financial burden on patients and the facility. Addressing these systemic barriers is essential for improving CM management and reducing its persistently high mortality in resource-limited settings.

A key limitation of this study is the reliance on data abstracted from patients’ medical records, which was constrained by missing information and the inability to seek clarification from prescribers when needed. We mitigated these risks by using a piloted abstraction tool, training and on-site supervision of abstractors, drawing from both paper charts and electronic systems, and applying logic checks during cleaning. We prespecified covariates, adjusted for hospital site and calendar period, excluded patients with unknown disposition from the primary analysis, and ran sensitivity analyses to test robustness. Residual confounding remains possible, particularly from unmeasured severity markers including mental status, intracranial pressure, CrAg titer and lack of time-to-event data. Furthermore, as a purely quantitative study, it does not capture the perspectives and lived experiences of HCPs and patients regarding the burden of CM, treatment practices, outcomes, and the challenges and opportunities that could be leveraged to improve management of this life-threatening opportunistic infection among PLHIV.

## V. Conclusion

This study demonstrates that CM remains a significant opportunistic infection among PLHIV, with most patients still managed with fluconazole monotherapy despite national and international guideline recommendations. Patients treated with amphotericin B–based regimens had a markedly higher likelihood of survival at discharge, underscoring the urgent need to align clinical practice with evidence-based guidelines. We therefore recommend that CM in PLHIV be managed using amphotericin B–based regimens as directed by local and WHO protocols. Additionally, future qualitative research should explore the barriers faced by HCPs, patients, and policymakers in implementing guideline-recommended CM treatment, to inform strategies that can strengthen CM management and reduce associated mortality.

## Data Availability

All relevant data are within the manuscript and its Supporting Information files.

## VI. List of Abbreviation

AIDS: acquired immunodeficiency syndrome
aPR: adjusted prevalence ratio
aRR: adjusted risk ratio
ART: antiretroviral therapy
CI: confidence interval
CM: cryptococcal meningitis
CrAg: cryptococcal antigen
cPR: crude prevalence ratio
CSF: cerebrospinal fluid
CTC: care and treatment centre
DRRH: Dodoma Regional Referral Hospital
HCPs: health care providers
HIV: human immunodeficiency virus
LTFU: lost to follow-up
MUHAS: Muhimbili University of Health and Allied Sciences
PLHIV: people living with HIV
RRHs: regional referral hospitals
SSA: sub-Saharan Africa
SPSS: Statistical Package for the Social Sciences
SRRH: Singida Regional Referral Hospital
WHO: World Health Organization.

## VII. Acknowledgement

The authors would like to thank the Dodoma and Singida Regional Referral Hospital administrations for issuing permission for data collection, the staff and research assistants for their support during data abstraction. We further acknowledge all patients whose redacted data made this study possible.

## Notes

### Competing Interest Statement

The authors have declared no competing interest.

### Funding Statement

The author(s) received no specific funding for this work.

